# Inward and outward effectiveness of cloth masks, a surgical mask, and a face shield

**DOI:** 10.1101/2020.11.18.20233353

**Authors:** Jin Pan, Charbel Harb, Weinan Leng, Linsey C. Marr

**Affiliations:** Civil and Environmental Engineering, Virginia Tech, Blacksburg, VA 24061

**Keywords:** masks, aerosol, transmission, COVID-19, SARS-CoV-2, face coverings

## Abstract

We evaluated the effectiveness of 11 face coverings for material filtration efficiency, inward protection efficiency on a manikin, and outward protection efficiency on a manikin. At the most penetrating particle size, the vacuum bag, microfiber cloth, and surgical mask had material filtration efficiencies >50%, while the other materials had much lower filtration efficiencies. However, these efficiencies increased rapidly with particle size, and many materials had efficiencies >50% at 2 μm and >75% at 5 μm. The vacuum bag performed best, with efficiencies of 54-96% for all three metrics, depending on particle size. The thin acrylic and face shield performed worst. Inward protection efficiency and outward protection efficiency were similar for many masks; the two efficiencies diverged for stiffer materials and those worn more loosely (e.g., bandana) or more tightly (e.g., wrapped around the head) compared to a standard earloop mask. Discrepancies between material filtration efficiency and inward/outward protection efficiency indicated that the fit of the mask was important. We calculated that the particle size most likely to deposit in the respiratory tract when wearing a mask is ∼2 μm. Based on these findings, we recommend a three-layer mask consisting of outer layers of a flexible, tightly woven fabric and an inner layer consisting of a material designed to filter out particles. This combination should produce an overall efficiency of >70% at the most penetrating particle size and >90% for particles 1 μm and larger if the mask fits well.

## Introduction

Amid mounting evidence that COVID-19 is transmitted via inhalation of virus-laden aerosols (Allen and Marr 2020; Asadi et al. 2020; Hadei et al. 2020; Morawska et al. 2020; Prather, Wang and Schooley 2020), universal masking has emerged as one of a suite of intervention strategies for reducing community transmission of the disease. There is a correlation between widespread mask wearing (The Economist 2020), or at least interest in masks (Wong et al. 2020), and lower incidence of COVID-19 by country and between mask mandates and county-level COVID-19 growth rates in the US (Lyu and Wehby 2020), but a causal relationship has not been confirmed.

Due to a shortage of medical masks and respirators, some public health agencies have recommended the use of cloth face coverings. While there have been numerous studies on the ability of surgical masks and N95 respirators to filter out particles, far less is known about the ability of cloth masks to provide both inward protection to reduce the wearer’s exposure and outward protection for source control. Ideally, a randomized controlled trial would be conducted, but in the absence of such evidence, we can evaluate the ability of masks to block particles under controlled conditions.

Reviews on the use of masks in both healthcare and non-healthcare settings to reduce transmission of other respiratory diseases mostly show a protective effect. A systematic review and meta-analysis of interventions against respiratory viruses found that wearing simple masks was highly effective at reducing transmission of severe acute respiratory syndrome (SARS) in five case control studies (Jefferson et al. 2008). In contrast, a review of 10 randomized controlled trials of mask wearing in non-healthcare settings concluded that there was not a substantial effect on influenza transmission in terms of risk ratio, although most of the studies were underpowered and compliance was not perfect (Xiao et al. 2020). A systematic review of interventions against SARS-CoV-2 and the coronaviruses that cause SARS and Middle East respiratory syndrome found that the use of face masks could result in a large reduction in the risk of infection (Chu et al. 2020).

Laboratory studies have demonstrated the ability of surgical masks to provide both inward and outward protection against viruses. Testing of eight different surgical masks on a manikin with influenza virus in droplets/aerosols of size 1–200 µm found that the amount of virus detected behind the mask was reduced by an average of 83%, with a range of 9% to 98% (Makison Booth et al. 2013). The ability of a mask to block influenza virus was correlated with its ability to block droplets/aerosols containing only phosphate buffered saline (PBS) and bovine serum albumin (BSA). Surgical masks used for source control on influenza patients during breathing and coughing reduced the amount of virus released into the air in coarse (> 5 µm) and fine (≤ 5 µm) aerosols by 96% and 64%, respectively (Milton et al. 2013). In a follow-up study, surgical masks blocked the release of seasonal coronaviruses in coarse and fine aerosols to undetectable levels, while they blocked influenza virus in most but not all patients (Leung et al. 2020).

There have been some studies of cloth masks, which have been found to be less protective than surgical masks in most, but not all, cases. A variety of cloth materials mounted in a filter holder removed 49% to 86% of aerosolized bacteriophage MS2, compared to 89% removal by a surgical mask (Makison Booth et al. 2013). According to fit tests on 21 adults in the same study, homemade, 100% cotton masks provided median inward filtration efficiencies of 50%, compared to 80% for surgical masks. The filtration efficiencies of 44 materials and medical masks, challenged with sodium chloride (NaCl) particles of diameter 0.03–0.25 μm, ranged from <10% for polyurethane foam to nearly 100% for a vacuum cleaner bag (Drewnick et al. 2020). Cloth masks, sweatshirts, t-shirts, towels, and scarves evaluated in a TSI Automated Filter Tester had filtration efficiencies of 10–60% against polydisperse NaCl particles ranging in size from 0.02 to 1.0 µm; the towels performed best (Rengasamy, Eimer and Shaffer 2010). Homemade masks made from tea cloths and worn by volunteers had a median inward filtration efficiency of 60%, compared to 76% for a surgical mask (van der Sande, Teunis and Sabel 2008). Pieces of a bandana, veil, shawl, handkerchief, and cotton t-shirt mounted in a filter holder and challenged with volcanic ash particles were found to have filtration efficiencies of 18% to 43% in terms of mass concentration (Mueller et al. 2018).

N95 respirators and cloth masks serve different purposes, so the testing procedure for N95s is not necessarily well-suited for cloth masks. An N95 must be able to protect an individual worker in high-risk situations. A critical component of its efficacy is the fit test to ensure that the respirator seals completely to the face with no leaks. On the other hand, the overall goal of wearing cloth masks during the COVID-19 pandemic is to reduce community transmission. Cloth masks provide some degree of both source control and exposure reduction. While an N95 must block at least 95% of NaCl particles of the most penetrating size, 0.3 μm, cloth masks can be effective if they remove at least some particles, particularly those of the size that is most relevant for transmission. Although we do not yet know which size particles are most important, we can make some inferences from existing studies. SARS-CoV-2 and other viruses are carried by particles ranging in size from <1 μm to >5 μm (Chia et al. 2020; Liu et al. 2020; Yan et al. 2018; Yang, Elankumaran and Marr 2011). A SARS-CoV-2 virion is 0.1 μm in diameter, but it is carried in respiratory droplets that also contain salts, proteins, and other components of respiratory fluid. Even if all the water evaporates, the mass of the non-volatile components is expected to be orders of magnitude larger than that of any viruses that might be present (Marr et al. 2019), so the size of a particle carrying an intact virus must be quite a bit larger than 0.1 μm. The smaller mode of respiratory particles produced during breathing and speaking is centered around 1 μm, and there are relatively few particles smaller than 0.5 μm (Johnson et al. 2011). Influenza transmission between ferrets has been shown to be mediated by particles larger than 1.5 μm (Zhou et al. 2018). Thus, it seems prudent to evaluate mask performance over a range of particle sizes, particularly those larger than 0.3 μm.

Given the advice of public health agencies for the general public to wear face coverings and the paucity of knowledge about their effectiveness, the objective of this study is to evaluate the efficiency of cloth masks compared to a surgical mask and a face shield at blocking particles over a wide range of sizes. We first measure the filtration efficiency of materials under ideal conditions and then investigate both inward and outward protection efficiency of the materials when worn as masks on a manikin. We expect that efficiency on a manikin will be lower than in a filter holder due to leakage around the mask and that outward efficiency will be higher than inward efficiency due to differences in velocity of the particles as they approach the material. The results of this study will contribute to understanding how universal masking might reduce transmission of COVID-19 and other respiratory diseases.

## Methods

### Masks

We tested nine materials that were fashioned into masks, one surgical mask, and one face shield, shown in Figure 1. To make the masks, we cut materials into 15.5 cm × 10 cm rectangles and securely taped them to a frame tailored from a surgical mask, except for two designs that followed instructions from the US Centers for Disease Control and Prevention (CDC). These included a sewn mask made of two layers of a 200-thread count cotton pillowcase and a non-sewn mask cut from a cotton t-shirt (Centers for Disease Control and Prevention 2020). The instructions for the non-sewn mask used in this study have been supplanted with an updated design involving a large square of fabric and rubber bands. The surgical mask had a single layer and was advertised to meet ASTM level 1 specifications, which require ≥95% filtration efficiency of particles larger than 1 μm. We characterized the texture and structure of the masks using a scanning electron microscope (FEI Quanta 600 FEG). Because it is not possible to generate or characterize particles spanning a wide range of sizes with a single experimental setup, we designed several different protocols for testing masks, optimizing among different types of equipment and detection limits, as described below.

**Figure 1.**
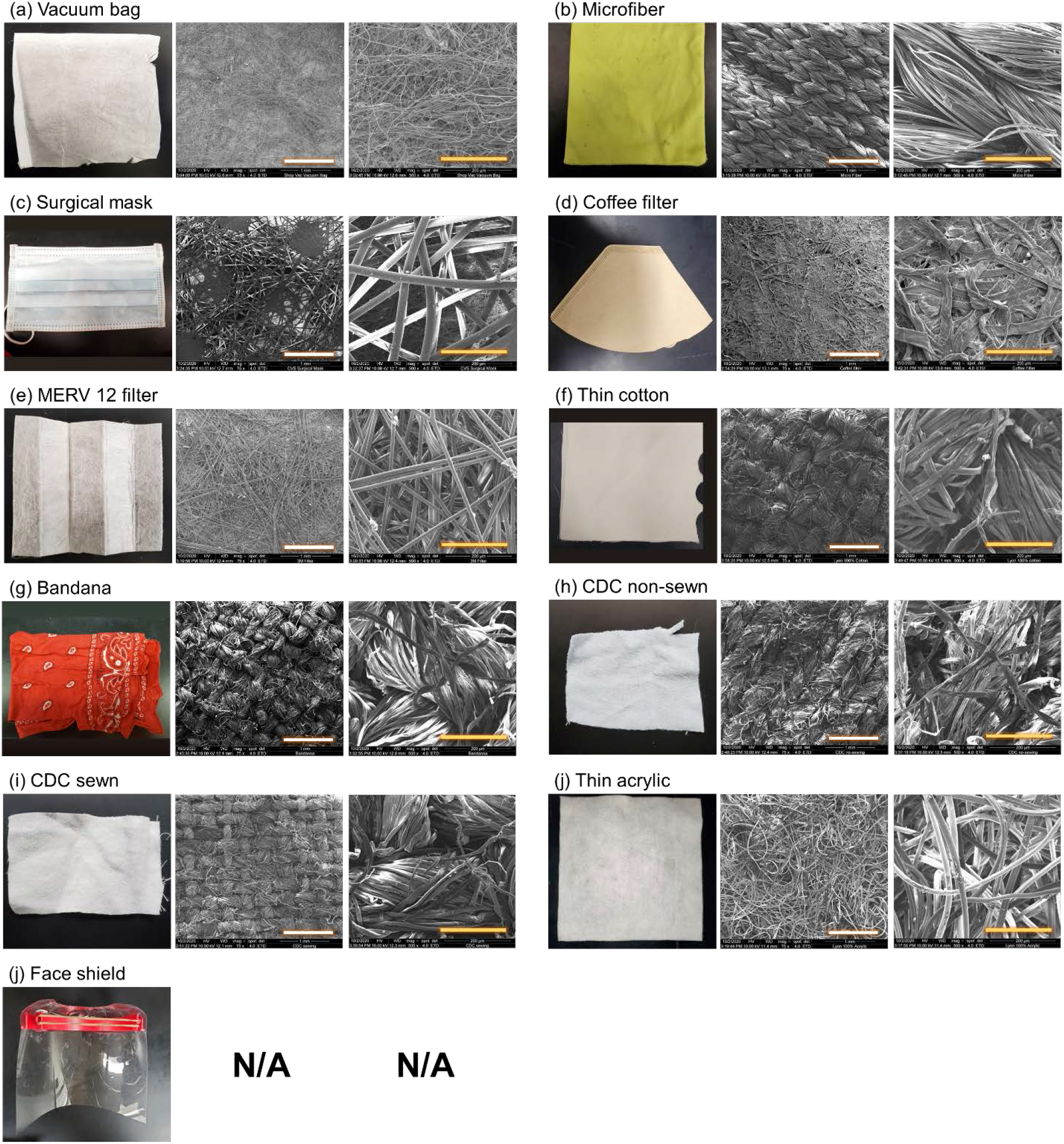
Ten mask materials and a face shield. SEM images are shown at two scales: the white scale bar represents 1 mm and the yellow one represents 200 μm. There are no SEM images for the face shield, which was made of a plastic sheet.

### Material filtration efficiency

Evaluation of the materials for filtration efficiency followed a protocol based on National Institute of Occupational Safety and Health (NIOSH) testing procedures. Using a Collison 3-jet nebulizer (BGI MRE-3, BGI Inc., MA, USA), we generated challenge particles of size 0.04 to 1 μm from a 2% NaCl solution. The particles filled a 280 L polyethylene chamber (Sigma AtmosBag, Sigma– Aldrich, ON, Canada), in which we placed a small fan to promote mixing. The temperature and humidity inside the chamber were 22 °C and 25-35% RH, respectively. We measured particle concentrations and size distributions using a scanning mobility particle sizer (SMPS 3936, TSI Inc., MN, USA), with the particle density set to 2.165 g/cm^3^ (NaCl) to convert from mobility diameter to aerodynamic diameter. We cut out circular pieces of each material to mount in a 25 mm stainless steel filter holder (Advantec, Cole Parmer, IL, USA) that was connected to a vacuum line whose flow rate was maintained at 2.7 L/min by a mass flow controller (32907-53, Cole Parmer, IL, USA). The SMPS sampled from this line at a rate of 0.3 L/min, producing a total flow rate of 3.0 L/min and a corresponding face velocity of 10 cm/s through the material. Clean make-up air flow to the chamber was provided through a high-efficiency particulate air filter capsule (12144, Pall Corporation, MA, USA). We checked the material filtration efficiency of an N95 respirator and the microfiber cloth with and without a Kr-85 radioactive neutralizer (3012, TSI Inc., MN, USA) or soft x-ray neutralizer (XRC-05, HCT CO., Ltd, Republic of Korea) after the nebulizer, and did not find significant differences (Figures S1-S3), so we did not employ a neutralizer in subsequent tests. To calculate the size-resolved filtration efficiency, we compared measurements with the material in the filter holder to those made with an empty filter holder, as shown in equation (1), where *FE* is the material filtration efficiency; *D*_*P*_ is the particle diameter; *C*_*blank*_ is the concentration of challenge particles measured downstream of the empty filter holder, and *C*_*material*_ is the concentration of particles downstream of the material:

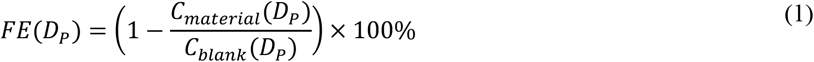

We conducted these experiments in triplicate using three different pieces cut from each material.

In addition to challenging the masks with submicron particles generated by the Collison nebulizer, we also tested larger particles ranging in size from 2 to 5 μm. We generated these from a 2% NaCl solution using a flow focusing monodisperse aerosol generator (FMAG, TSI Inc., MN, USA). We measured the particles using an aerodynamic particle sizer spectrometer (APS 3321, TSI Inc., MN, USA). Because the APS samples at a flow rate of 1.0 L/min, we adjusted the vacuum line to 2.0 L/min to produce a total flow rate of 3.0 L/min, the same as used for testing smaller particles. Clean make-up air was also applied as described above. We calculated the filtration efficiency according to equation (1) in triplicate. We also measured the pressure drop of each material in the filter holder using a differential pressure gauge (Minihelic II 2-5005, Dwyer Instruments, IN, USA).

### Inward and outward protection efficiency at close distance

We evaluated both inward and outward protection efficiency of face coverings using two manikins mounted on opposite sides of a 57-L acrylic chamber (51 cm × 34 cm × 33 cm), mimicking the situation of close talking, with a mouth-to-mouth distance of 33 cm (Figure 2a, b). The “exhaling” manikin was connected to a medical nebulizer (AIRIAL) filled with 2% NaCl solution, that produced a flow rate of 10 L/min through 0.79 cm i.d. tubing. The “inhaling” manikin was connected to both the APS and a vacuum line, with flow rates of 1 L/min and 14 L/min, respectively, resulting in a total flow rate of 15 L/min through 1 cm i.d. tubing. Make-up air entered the chamber around the top perimeter, to minimize disruption to air flow that might be introduced by a port, and had a background particle concentration of at most 0.5% of that generated in the chamber by the nebulizer. The air velocity at both manikin’s mouths was 3.2–3.4 m/s, similar to that of breathing and talking (Gupta, Lin and Chen 2010; Xie et al. 2009). To minimize losses of particles, we used conductive tubing in lengths as short as possible.

**Figure 2.**
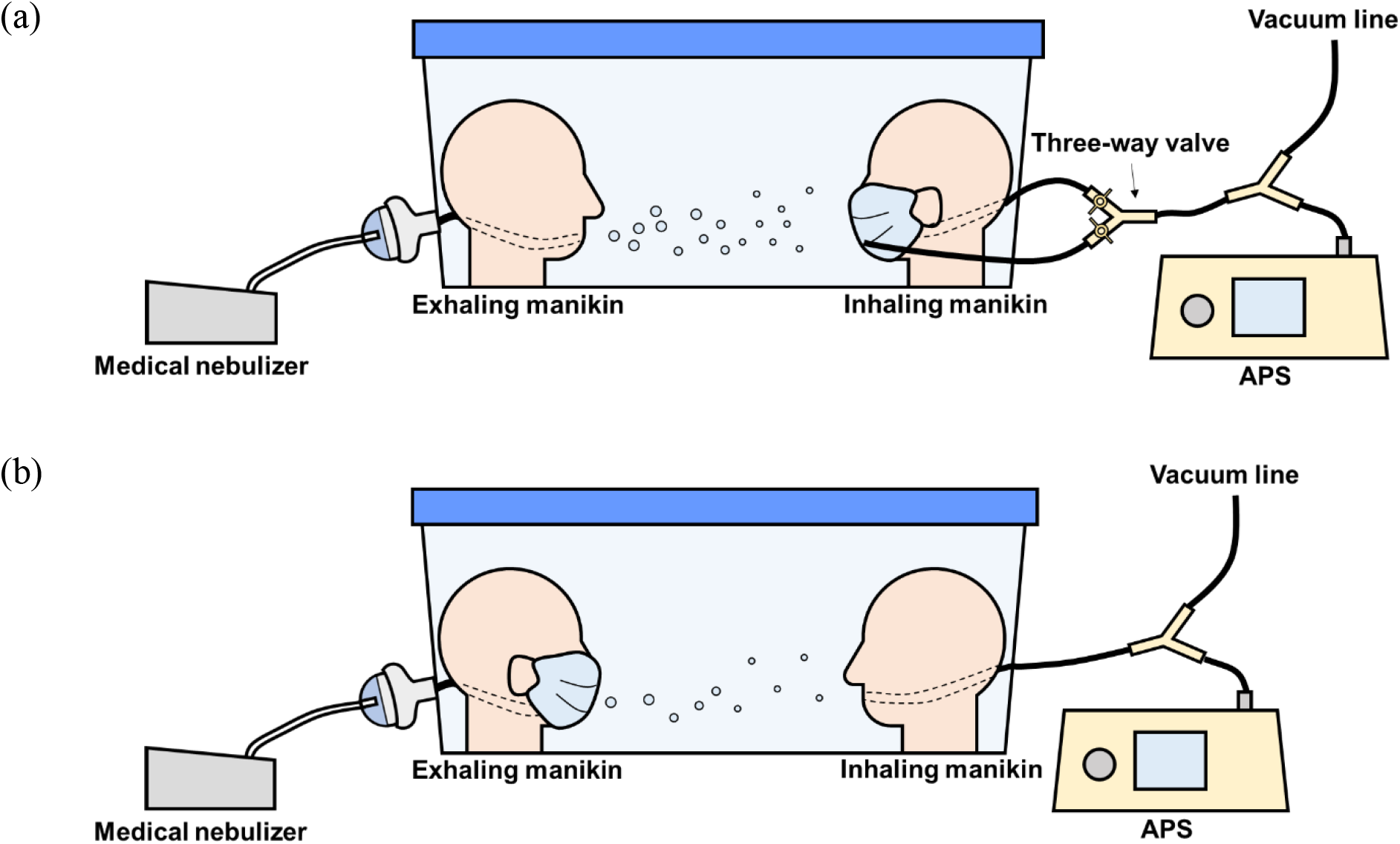
Schematic of experimental setup for determining (a) inward protection efficiency and (b) outward protection efficiency.

To evaluate inward protection efficiency, we attached face coverings to the inhaling manikin (Figure 2a, Figure S4) and tested two scenarios. In scenario 1, we ran the medical nebulizer for 3 s through the exhaling manikin, generating particles of size 0.5–2 μm. Using a three-way valve, we set up the APS to sample either through the inhaling manikin’s mouth or through tubing whose inlet was placed outside the face covering, near the manikin’s mouth. The flow rate through the inhaling manikin remained constant at 15 L/min. We then waited 30 s for particle concentrations to decay below the upper limit of detection of the APS, switched the valve to sample from outside the mask, and measured the size distribution in the chamber for 5 s, denoted *C*_*c1*_. We then switched the valve so that the APS sampled through the inhaling manikin’s mouth and measured particles that penetrated the mask, denoted *C*_*m*_. To account for the continually decaying particle concentration in the chamber, we then switched back to measuring particles in the chamber again, denoted as *C*_*c2*_. The difference between *C*_*c1*_ and *C*_*c2*_ was less than 10% in all cases. Therefore, we used the average of *C*_*c1*_ and *C*_*c2*_ to represent *C*_*c*_ at the time when we measured *C*_*m*_. We calculated the inward protection efficiency based on equation (1), replacing the numerator with *C*_*m*_*(D*_*P*_) and the denominator with *C*_*c*_*(D*_*P*_). The temperature and humidity inside the chamber were 22 °C and 50–70% RH, respectively. In a separate experiment, we demonstrated that the three-way valve and the location of sampling inlets did not bias the calculation. There was no difference in the concentration and size distribution of particles whether the APS sampled directly from the chamber or through the inhaling manikin without the mask. Measurements in scenario 2 followed a similar protocol as in scenario 1 except that the medical nebulizer ran for 30 s instead of 3 s to generate larger particles, up to 5 μm, thanks to coagulation.

To evaluate outward protection efficiency, we removed the three-way valve and connected the APS and vacuum line directly to the inhaling manikin (Figure 2b). In each test, we ran the medical nebulizer for 30 s and then allowed particle concentrations to decay, as in scenario 2 of the inward protection protocol, and we measured the chamber concentration (*C*_*c1*_) using the APS at 1-s resolution. After introducing the HEPA-filtered air to flush particles from the chamber, we then put the mask or face shield on the exhaling manikin and ran the medical nebulizer for 30 s again to measure the concentration (*C*_*m*_). Then we flushed the chamber again, ran the nebulizer to measure the chamber concentration *C*_*c2*_, and calculated the average *C*_*c*_ as described in scenario 1. We calculated the outward protection efficiency according to equation (1) as well. We conducted all measurements in triplicate.

### Droplet deposition analysis

We evaluated the ability of the face coverings to block droplets larger than 20 μm, which is the upper limit of the APS, using a modified droplet deposition analysis (DDA) (Johnson et al. 2011; Xie et al. 2009). The setup was similar to that of the outward protection protocol but with an air brush (MP290001AV, Campbell Hausfeld, OH, USA) in place of the medical nebulizer to generate larger droplets (Lindsley et al. 2013). We connected the air brush to HEPA-filtered air and a gas regulator set at 165.5 kPa, resulting in a total flow rate of 10 L/min, the same as the flow rate of the medical nebulizer. We filled the air brush with 2% NaCl solution and red food dye at a ratio of 4:1. We taped five glass slides (75 mm × 25 mm) to the face of the inhaling manikin. We pre-cleaned each slide using 70% isopropyl alcohol wipes.

First, we sprayed the air brush for 3 s without the face covering on the exhaling manikin. We then removed the glass slides from the inhaling manikin and inspected them under an optical microscope at 10× magnification (EVOS FL Auto, Life Technologies, CA, USA). We put the face covering on the exhaling manikin and repeated the same steps. To identify droplets on the slides, we processed the images using ImageJ and then manually counted the stains and measured their size with a limit of detection of 12.3 μm/pixel. Because the droplets spread upon impaction with the slides, we corrected their size assuming a spread factor of 1.5, the ratio of the size of the stain to the original diameter of the droplet (Johnson et al. 2011). We conducted all measurements in triplicate.

## Results

### Size of challenge particles

We used four different types of aerosol generators to cover a broad size range and to accommodate different setups. The Collison nebulizer and FMAG, used to determine material filtration efficiency, generated particles ranging in size from 0.04 to 1 μm and from 2 to 5 μm, respectively (Figure 3a, b). The Collison nebulizer produced particles with a geometric mean diameter (GMD) of 0.12 μm and geometric standard deviation (GSD) of 1.4, and the FMAG a GMD of 4 μm and GSD of 1.21. The figure also shows the size distribution measured downstream of a MERV 12 filter to illustrate the data used to calculate filtration and protection efficiencies. The medical nebulizer produced particles ranging in size from 0.5 to 5 μm; the GMD was below the detection limit of the APS (Figure 3c). As the medical nebulizer covered a relatively large size range, we chose to use it to evaluate the inward and outward protection efficiencies (Figure 3c). The air brush generated large particles ranging in size from 20 μm to greater than 135 μm (Figure 3d).

**Figure 3.**
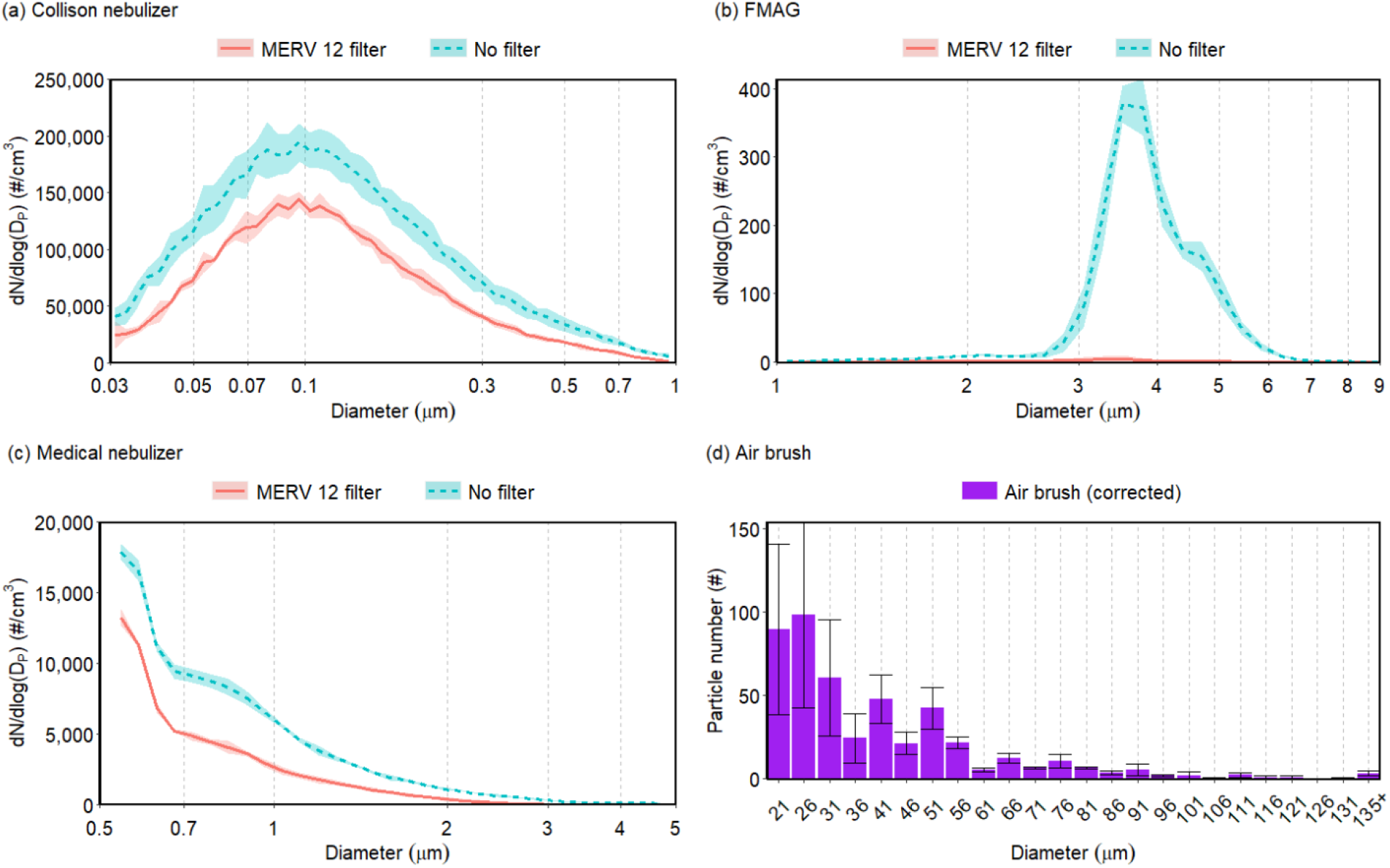
Concentration and size distribution of particles as a function of aerodynamic diameter generated by (a) Collison nebulizer, (b) FMAG, (c) medical nebulizer, without a filter (blue dashed line) or downstream of a MERV 12 filter (red solid line), and (d) air brush. In panel (d), the diameter was corrected from the measured size of the droplet stains on the slide by a factor of 1.5. Shading and error bars represent the standard deviations of triplicates.

### Material filtration efficiency

We tested the material filtration efficiency of nine common homemade mask materials and one surgical mask. We did not test the face shield because it does not allow air flow through it. Figure 4 shows results obtained using the Collison nebulizer and SMPS over the size range 0.04 to 1 μm. The efficiency curves exhibit the expected U shape with a minimum in most cases in the range 0.1–0.3 μm, where no collection mechanism is especially efficient (Hinds 1999).

**Figure 4.**
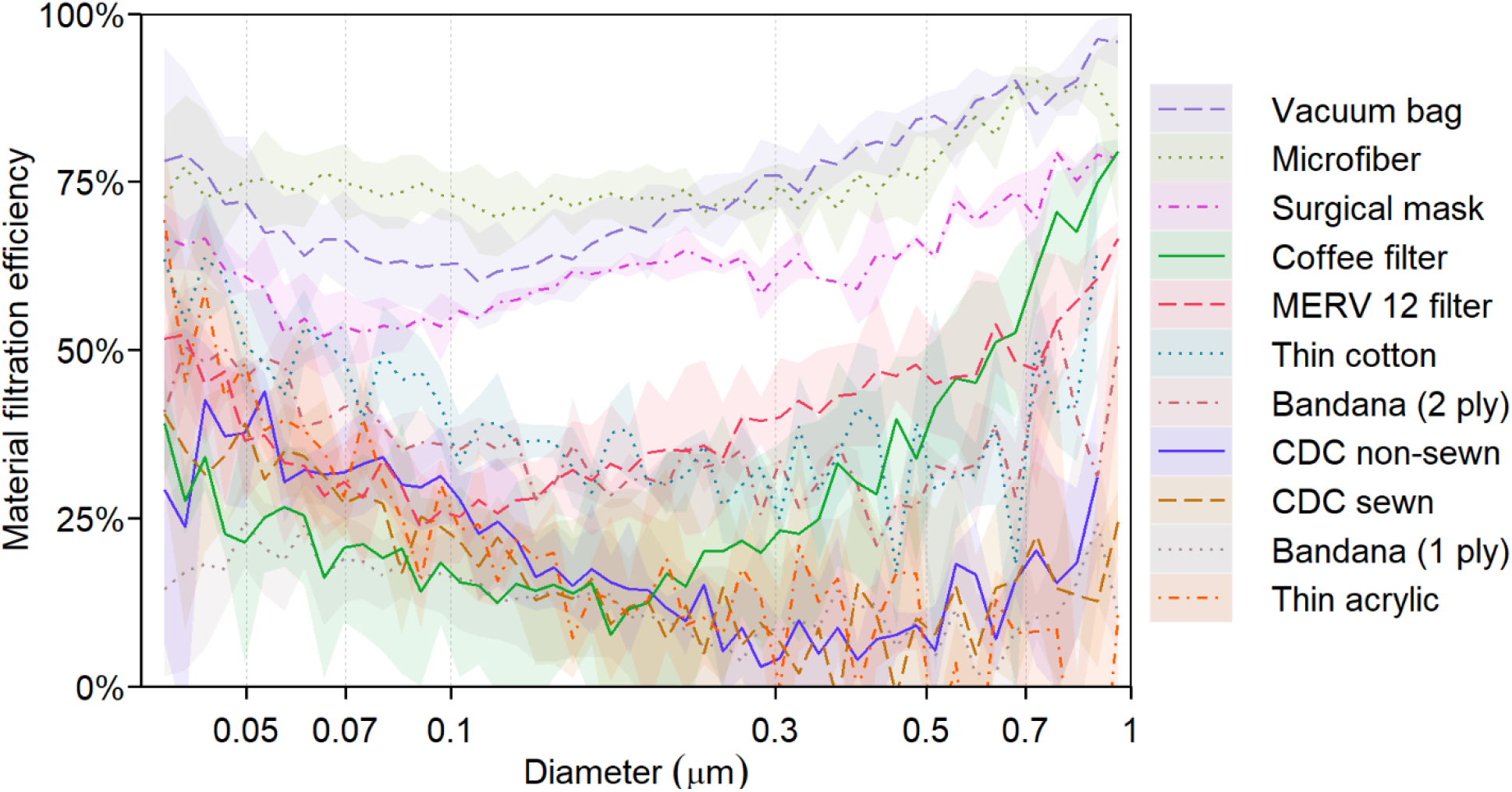
Material filtration efficiency of 10 mask materials as a function of aerodynamic diameter. The bandana appears twice because it was tested in both 1-ply and 2-ply configurations. Shading represents the standard deviations of triplicates.

The vacuum bag and microfiber performed best, with a minimum efficiency of 60%. Other studies have also reported that vacuum bags have high filtration efficiencies (Drewnick et al. 2020; Zangmeister et al. 2020), whereas the performance of microfiber varies depending on the manufacturer and fabric structure (Drewnick et al. 2020; Zhao et al. 2020). Following the top two, the surgical mask was ∼50–75% efficient over this size range, falling with the range reported for surgical masks in previous studies (Makison Booth et al. 2013; Oberg and Brosseau 2008; Zangmeister et al. 2020; Zhao et al. 2020). The minimum efficiency of the coffee filter was only 10% for particles at 0.17 μm, lower than the reported value of 34.4% in another study (at a face velocity of 6.3 cm/s) (Zangmeister et al. 2020), but its efficiency rapidly increased with particle size to 75% for particles at 1 μm. The MERV 12 filter reached its lowest efficiency of 25% at 0.1 μm and had an efficiency > 50% at the extremes shown in Figure 4. Common fabrics, including the thin cotton and bandana (2 ply), had low efficiencies, mostly between 30% and 50%. The fabrics fashioned into the CDC non-sewn and CDC sewn masks, bandana (1 ply), and thin acrylic had even lower efficiencies of 5–40% for submicron particles.

Most of the materials exhibited a much better material filtration efficiency for particles >1 μm than for smaller ones, as shown by the black solid line in Figure 5. The vacuum bag, microfiber, surgical mask, and MERV 12 filter achieved 90% or higher efficiency at 2 μm, and thin cotton and coffee filter were around 80% efficient at this size. The 2-ply bandana performed much better than the 1-ply bandana, with efficiencies of ∼75% and <40% at 2 μm, respectively. The CDC non-sewn and CDC sewn mask materials had efficiencies of ∼50% at 2 μm, and their efficiencies increased with particle size to up to 75% at 5 μm. The thin acrylic still ranked at the bottom. Its efficiency was <30% at 2 μm but reached 75% at 5 μm.

**Figure 5.**
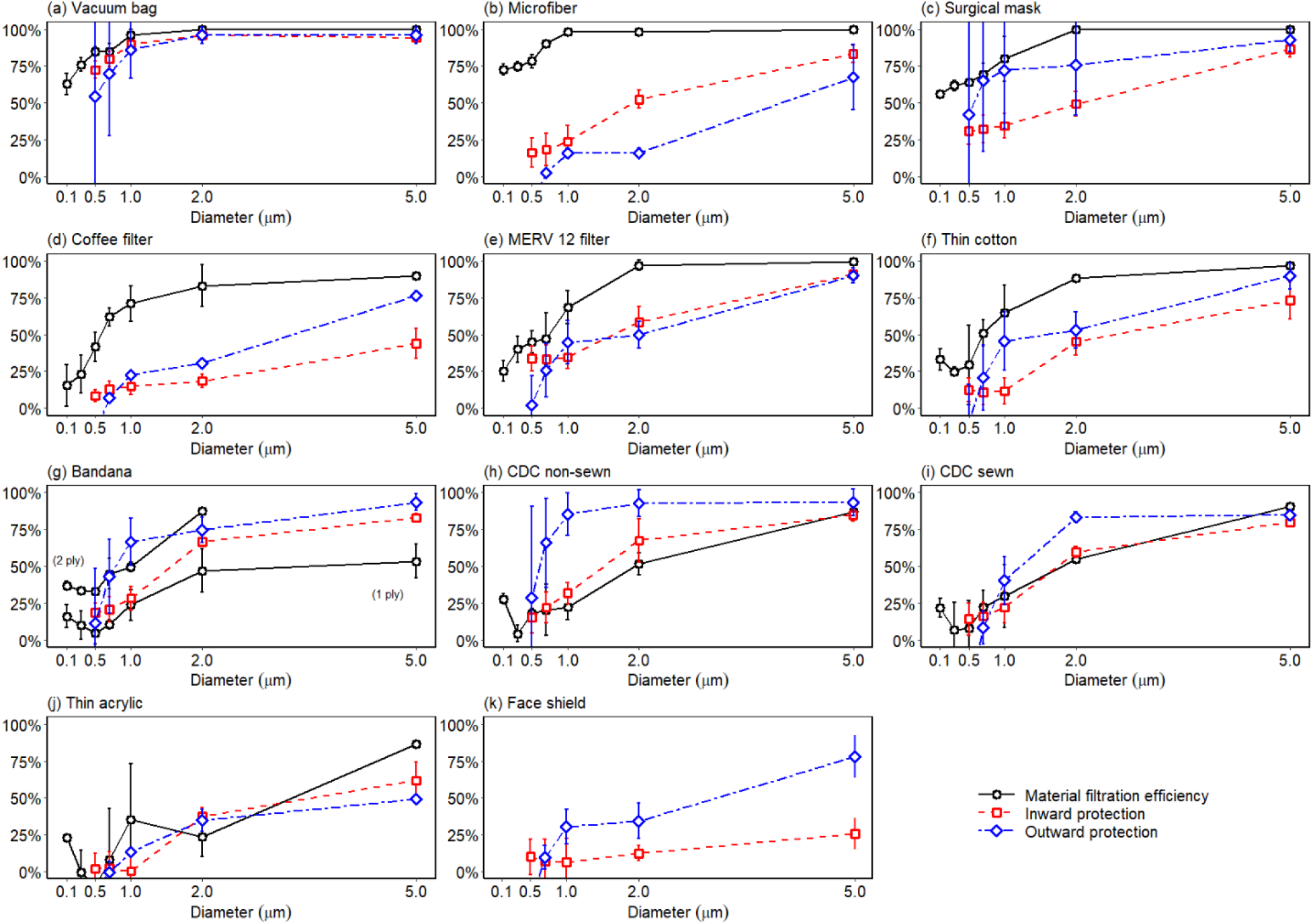
Inward and outward protection efficiency of 10 masks and a face shield. The face shield was not tested for material filtration efficiency because it did not allow air flow through the material. Error bars represent the standard deviations of triplicates.

Figure 6 shows the pressure drop across all materials, measured at a flow rate of 3.0 L/min through a sample 25 mm in diameter. Microfiber had the highest pressure drop, nearly 1000 Pa, followed by the coffee filter and thin cotton at ∼380 Pa. The pressure drop through the other materials was <150 Pa, among which the thin acrylic and MERV 12 filter had the lowest values, ∼70 Pa. We further related the pressure drop to the material filtration efficiency using a filter quality factor (*Q*), as defined by equation (2) (Hinds 1999; Podgórski, Bałazy and Gradoń 2006), where *FE(D*_*P*_) is the material filtration efficiency at a particle size of *D*_*P*_, and Δ*P* is the pressure drop:

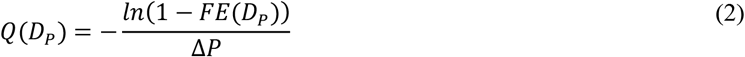

**Figure 6.**
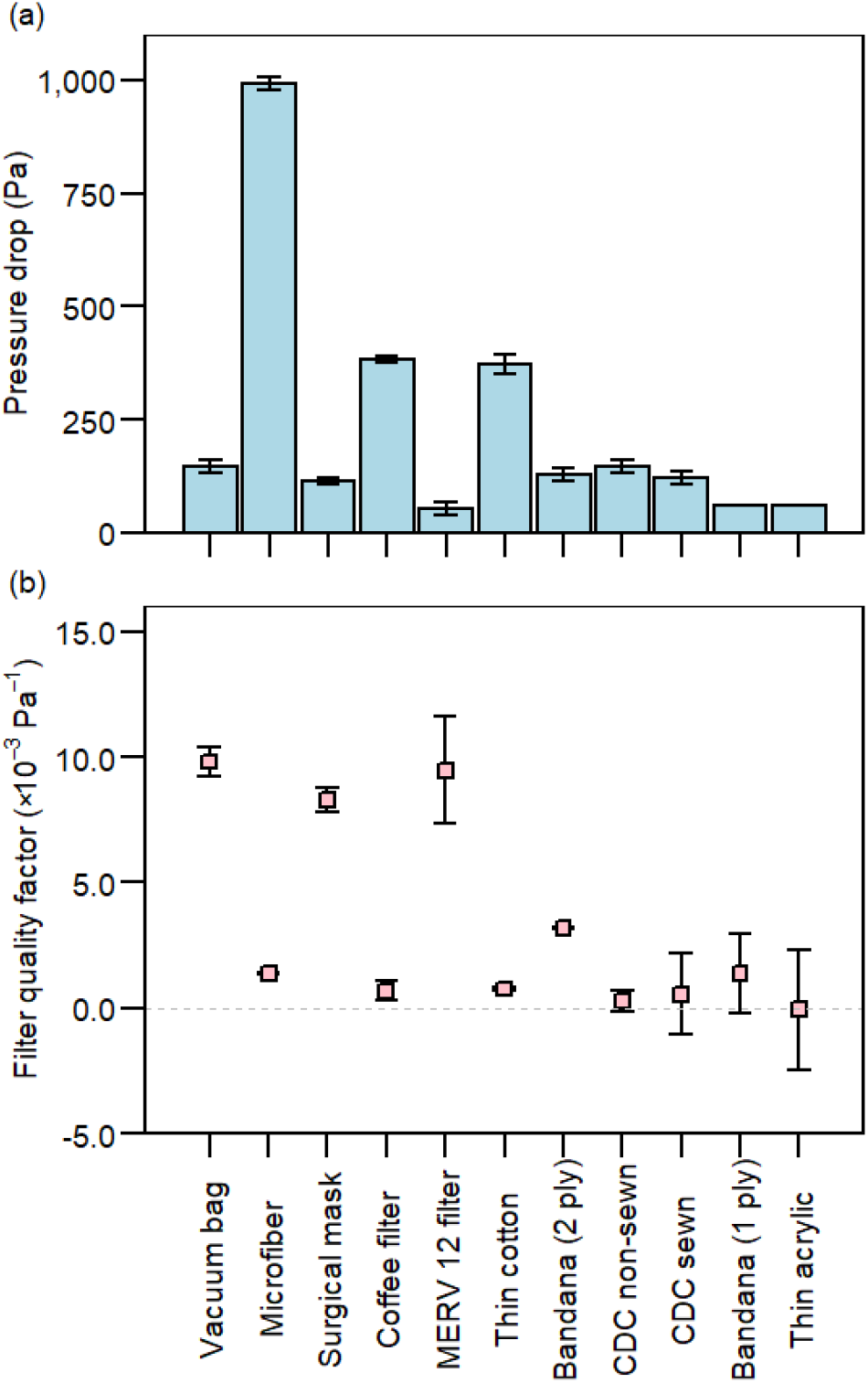
Pressure drop and filter quality factor at 0.3 μm of 10 mask materials, sorted on the basis of material filtration efficiency (Figure 4). The bandana appears twice because it was tested in both 1-ply and 2-ply configurations. Error bars represent standard deviations of triplicates. In panel (a), there are no error bars for the last two materials as the measurements fell below the detection limit of the pressure gauge.

We chose 0.3 μm as the representative particle size for the calculation of *Q* for ease of comparison with other studies (Figure 6b). Since pressure drop is directly correlated with the breathability of the material, a high *Q* means a high filtration efficiency can be achieved with a low pressure drop, indicating that the material is efficient and easy to breathe through. The vacuum bag and MERV 12 filter, which are both designed to filter out particles, had the highest *Q* of all the materials (∼10×10^−3^ Pa^-1^). The surgical mask also performed well, with an average *Q* of 7.6×10^−3^ Pa^-1^, not significantly different from *Q* of the previous two. These results are comparable to those reported in another study conducted under similar conditions (Zangmeister et al. 2020). The *Q* values of the thin cotton, bandana (2 ply), and the other fabrics were <5×10^−3^ Pa^-1^, similar to previously reported values (Zangmeister et al. 2020; Zhao et al. 2020). Notably, an increase in the number of layers of the bandana resulted in an increase in *Q*.

SEM images of the materials’ structure can partly explain the differences in the performance. The vacuum bag, which had the highest material filtration efficiency, had the smallest-diameter fibers and such a compact structure that the pores or intervals between fibers were the least perceptible among all the materials (Figure 1a). The fibers of the microfiber cloth were also more tightly woven than those of other materials (Figure 1b), resulting in good filtration efficiency. The materials with low efficiency were generally loosely woven, such as the bandana (1 ply), 200-thread-count pillow case used for the CDC non-sewn mask, cotton t-shirt used for the CDC sewn mask, and thin acrylic (Figure 1g-j). However, the tightness of the weave was not the only factor influencing the filtration efficiency. For example, the fiber intervals were large for the surgical mask, yet it was composed of multiple layers of different materials (Zhao et al. 2020), which made it unique from other materials. That fabric structure alone does not explain filtration efficiency also applies to the filter quality factors. For instance, the vacuum bag had a compact texture yet a low pressure drop, resulting in a high *Q* value. Likewise, the surgical mask was not tightly woven, but it was more efficient and thus had a higher *Q* than many other materials. The number of layers (Drewnick et al. 2020), the properties of the fibers including diameter and electrostatic charges (Konda et al. 2020; Ou et al. 2020; Podgórski, Bałazy and Gradoń 2006; Zangmeister et al. 2020), and the material composition (Zangmeister et al. 2020; Zhao et al. 2020) all contribute to differences in filter quality factors.

### Inward and outward protection efficiency

In this study, the inward protection efficiency (*IPE*) quantifies the capability of a mask, as worn on a manikin, to protect the wearer by filtering out particles moving in the inward direction through the mask, from the surrounding air to the wearer’s respiratory tract. The outward protection efficiency (*OPE*) quantifies the capability of a mask for source control, to filter out particles moving in the outward direction through the mask, from the wearer to the surrounding air. After being made into a mask, the vacuum bag still ranked first for protection efficiency in both directions, with its *IPE* and *OPE* curves close to the material filtration efficiency curve (Figure 5a), especially for particles larger than 1 μm. Both *IPE* and *OPE* were >50% at 0.5 μm and >90% for particles larger than 2 μm. However, there were large variations in *OPE* for particles smaller than 0.7 μm. The *IPE* and *OPE* were also similar to the respective material filtration efficiency for the CDC-sewn and thin acrylic masks (Figure 5i, j), though their performance was much worse than that of the vacuum bag. The *OPEs* of the CDC sewn mask and thin acrylic mask were ∼75% and ∼50%, respectively, for particles larger than 2 μm, and both masks were not effective at blocking particles smaller than 0.7 μm. Notably, the *OPE* of the CDC sewn mask was slightly higher than its *IPE* at 2.0 μm, whereas no significant differences (*p*>0.05) between *OPE* and *IPE* were observed across all sizes for the thin acrylic mask.

In contrast, the microfiber and coffee filter masks had a much worse *IPE* and *OPE* than their material filtration efficiency (Figure 5b, d), indicating leakage and a poor fit. The *OPE* for the microfiber mask was <25% for particles smaller than 2 μm, a difference of >50 percentage points compared to its material filtration efficiency. Its *IPE* was slightly better but still 20–50 percentage points lower than its material filtration efficiency for particles smaller than 2 μm. Similar trends were also observed for the coffee filter, except that its *OPE* was slightly higher than its *IPE* at particle sizes larger than 2 μm.

For the surgical mask, thin cotton, and MERV 12 filter, the differences between *OPE* or *IPE* and material filtration efficiency were moderate, usually within 25 percentage points (Figure 5c, e, f). The *OPEs* of the surgical mask and thin cotton mask were higher than their *IPEs* but not significantly; and these efficiencies were lower than the corresponding material filtration efficiency. In particular, the average *OPE* of the surgical mask was substantially better than its *IPE* at particle sizes ranging from 0.7 to 2 μm, but given the large variability in *OPE*, such differences were not statistically significant (*p*>0.05). There were no significant differences (*p*>0.05) between *IPE* and *OPE* for the MERV 12 filter across all sizes.

The bandana, CDC non-sewn mask, and the face shield had unique forms. The bandana was folded in half in a triangle to mimic how people would normally wear it; its *IPE* and *OPE* fell in between the single-layered and double-layered material filtration efficiency (Figure 5g), with the *OPE* higher than *IPE* at a particle size of 1 μm (*p*<0.05). The CDC non-sewn mask, whose fit can be adjusted by tightening or loosening the straps, had an *OPE* that was significantly (*p*<0.05) higher than the material filtration efficiency at sizes ranging from 1 to 2 μm. It is likely that stretching or loosening the fabric altered its filtration efficiency. Its average *OPE* was also higher than the *IPE*, whereas no significant difference was found between its *IPE* and material filtration efficiency. The face shield did not block almost any aerosols smaller than 0.7 μm, as expected, for it did not fit closely to the manikin and thus allowed virus-laden aerosols to travel freely around the shield. However, it exhibited a decent *OPE* for particles at 5 μm (∼75%) and an *IPE* of ∼25% for such particles.

Figure 7 compares the *IPE* and *OPE* across all masks. The vacuum bag mask had the best performance in both directions, while the coffee filter mask, thin acrylic mask, and face shield ranked at the bottom. The CDC non-sewn mask and surgical mask followed the vacuum bag closely for *OPE* but not *IPE*. Interestingly, the *OPE* values for masks tested spanned a wide range, whereas their *IPE* values were closer, except for the vacuum bag. In addition, direct comparison of the two panels in Figure 7 reveals that *OPE* tended to be higher than *IPE*, illustrating that many face coverings work better for source control than protection of the wearer, although the difference was not significant in most cases.

**Figure 7.**
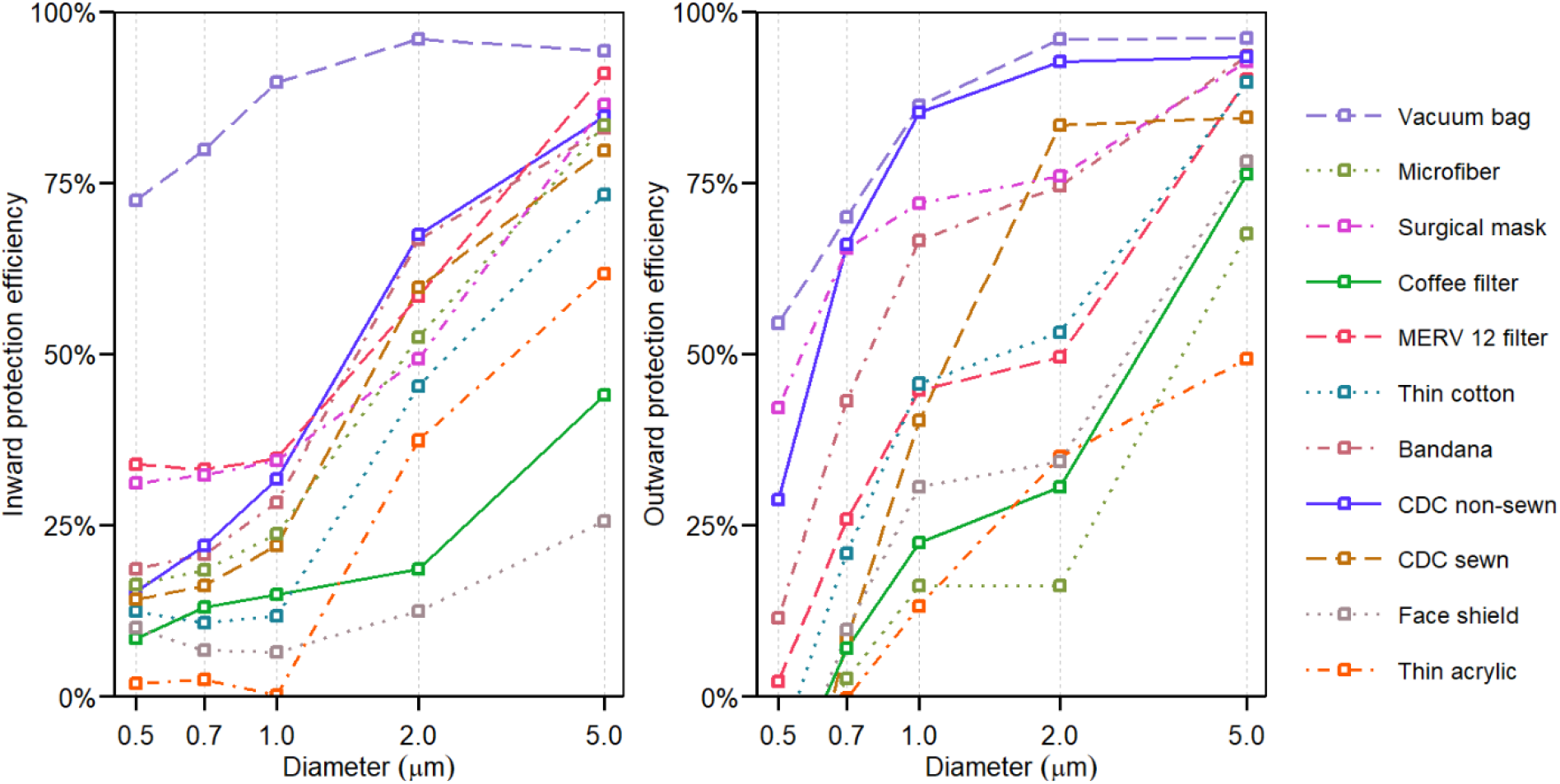
Inward and outward protection efficiency for all masks. For improved readability, error bars are not shown here, but they appear in Figure 5.

In response to a study that suggested that neck gaiters offer very little protection (Fischer et al. 2020), we measured the *OPE* of two neck gaiters, one made of thin 100% polyester and another made of a double layer of microfiber fabric that was 87% polyester and 13% elastane. Their average *OPEs* were at least 50% at 1 μm and >90% at 5 μm (Figure S5, S7), similar to the results for the CDC non-sewn mask. When doubled over, the thin polyester neck gaiter achieved an *OPE* of >90% over the size range of 0.5–5 μm (Figure S6). Due to the late addition of these face coverings, we were not able to measure their material filtration efficiency or *IPE*.

Droplet deposition analysis found no stains in the slides for all face coverings, indicating that all of them were able to prevent droplets larger than 20 μm from spreading 33 cm away.

## Discussion

For most of the face coverings tested, those with a high material filtration efficiency also had a better *OPE* and *IPE*. One example is the vacuum bag, which achieved outstanding performance compared to other materials with regards to material filtration efficiency, *IPE*, and *OPE*. It was able to filter out at least 60% of particles under perfect conditions and had an *OPE* and *IPE* of at least 50% and 75%, respectively, for particles 0.5 μm and larger. The MERV 12 filter, surgical mask, thin cotton, and CDC sewn mask also had decent material filtration efficiencies, *OPEs*, and *IPEs*, whereas the thin acrylic mask performed worst or near-worst on all three metrics. However, there were some exceptions, such as the microfiber cloth and coffee filter. The material filtration efficiencies of these two masks was much higher than their *OPEs* and *IPEs* (Figure 5b, d). The coffee filter and microfiber were thick and stiff, resulting in a poor fit with larger gaps between the manikin and the mask, through which particles could short circuit the mask. In contrast, the vacuum bag was thin and soft, which allowed it to conform to the face easily and achieve a high *IPE* and *OPE*. Hence, we propose that the stiffness of the material impacts the fit of the mask and can be responsible for large discrepancies between the material filtration efficiency and *OPE* and *IPE*. Additionally, differences in mask use among individuals will lead to variability in fit and thus effectiveness.

The CDC non-sewn mask was another exception. Generally, the *IPE* or *OPE* should be lower than the material filtration efficiency because the latter was tested in a filter holder with no opportunity for leaks. Nonetheless, the CDC non-sewn mask had a higher *OPE* than its material filtration efficiency. This unexpected result may be due to its unique form, resulting in a different way of it being stretched. Its two straps can be adjusted to fit it more tightly to the manikin face, especially to the mouth opening. Hence, the increased pressure caused by the expiratory flow was not able to push the CDC non-sewn mask outwards to create gaps between masks and the manikin like other conventional masks do (Lei et al. 2013; Liu et al. 1993; Mittal, Ni and Seo 2020), minimizing air leakage and bypass through the gaps. The stretching of the fabric may have caused a change in pore size and woven structure, which further impacted the filtration efficiency. In addition, the masks themselves also reduced the expired air velocity, which caused the particles to deposit before they could reach the sampling device, as shown in other studies (Hsiao et al. 2020; Mittal, Ni and Seo 2020; Tang et al. 2009). The combined effects of reduced gaps and reduced air velocity resulted in a uniquely high *OPE* for the CDC non-sewn mask. For other masks with a conventional shape, however, these two effects seemed compensatory during evaluation of *OPE*. While the masks caused a decrease in the expiratory air velocity, they were also pushed outwards by the outgoing flow, creating larger gaps between the masks and manikin. The contradiction in part explained why the differences between *OPE* and *IPE* were not as large as expected for the masks with conventional shapes, and why the bandana achieved an *OPE* better than expected, because it created a larger plenum between itself and the manikin that provided additional containment of the flow to lower the pressure drop and slow the flow jets through the gaps.

During the testing of *IPE*, we noticed that the vacuum through the inhaling manikin can suck the mask tightly against inlet opening, thus reducing the size of any gaps. This can explain the small differences between the material filtration efficiency and *IPE*, except for the coffee filter and microfiber as they were stiff and hard to move. However, this phenomenon also illustrates the tradeoff between breathability and filtration efficiency. Therefore, it is important to select fabrics that can achieve both high filtration efficiency and low pressure drop for making masks.

We also observed variable hydrophobicity of the mask material during the testing of *IPE* and *OPE*. The fabrics (e.g., thin cotton and thin acrylic) and coffee filter were wetted easily by droplets, whereas the filter materials, including the vacuum bag and the MERV 12 filter, were hydrophobic and kept the droplets on the surface of the material. El-Atab et al. developed a reusable hydrophobic mask and proposed that the hydrophobicity of the mask material might contribute to repelling the droplets (El-Atab et al. 2020). However, the role of hydrophobicity in filtration efficiency, *IPE*, and *OPE* remains unclear.

Whether particles actually deposit along the respiratory tract, potentially delivering an inhaled pathogen to a receptor, depends on two factors: (1) their ability to be inhaled into the respiratory tract and (2) their likelihood of depositing. The first can be reduced by a mask, and the second can be predicted as a function of particle size. Accounting for these two factors, we calculated the masked deposition rate (*MD*) by combining the inward protection effectiveness (*IPE*) and the International Commission and Radiological Protection (ICRP) model (Hinds 1999), as shown in equation (3):

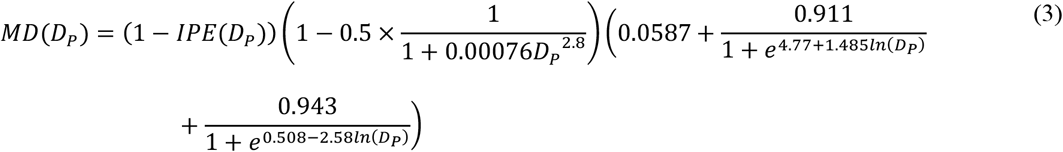

Figure 8 shows the masked deposition rate as a function of particle size. Here, lower values are better. The vacuum bag performed best, with a deposition rate of <10% across all sizes. The thin acrylic mask, the coffee filter mask, and the face shield were the worst, with a 50% or higher deposition rate at a particle size of 2 μm. Although there is considerable concern about exposure to virus in the smaller particles, the particles with the highest deposition rate were those around 2 μm. For example, SARS-CoV-2 RNA has been detected in particles in the size range of 1–4 μm (Chia et al. 2020). The smallest particle size considered in this analysis was 0.5 μm, but the deposition efficiency of 0.3 μm particles in the respiratory tract is even lower, so it is possible that concerns about mask efficiency at this size are overstated.

**Figure 8.**
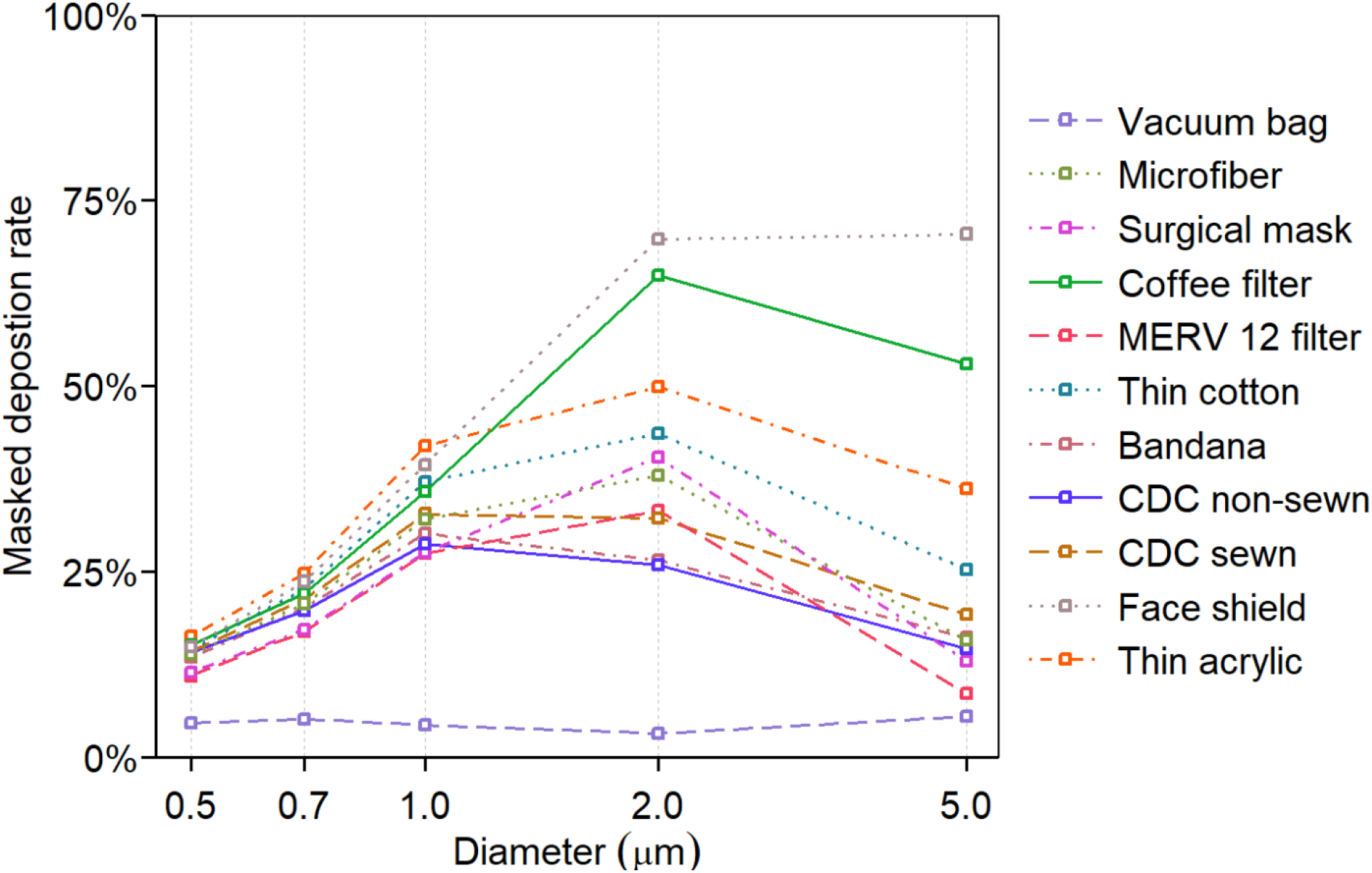
Masked deposition rate of 10 masks and a face shield as a function of the aerodynamic diameter.

This study was designed to test masks under tightly controlled conditions, which necessitate the use of mechanical particle generation and manikins instead of humans. However, this approach presents several limitations. The manikins are much more rigid than human skin, so masks may not fit as tightly. A study involving a head form with pliable, elastomeric skin found that fit factors of respirators were comparable to those measured on humans (Bergman et al. 2015), whereas in prior studies with head forms made of more rigid material, the fit factors were not as good (Bergman et al. 2014). In addition, our manikins did not perfectly mimic human respiratory activities because the aerosol flow traveled in only one direction in the inhaling manikin and the exhaling manikin. As discussed above, inhalation and exhalation will alter the plenum between the mask and the manikin, thus resulting in changes of the pressure drop and expiratory jets. We investigated only one flow rate out of the possible spectrum from gentle breathing to vigorous sneezing. Additionally, masks fit differently on different head shapes. Therefore, the performance of the masks on a human face under real-world conditions will certainly vary from the experimental results in this study. We did not test masks constructed of multiple layers of fabric, as prior work has shown that overall filtration efficiency is readily predicted by combining individual layers in series (Drewnick et al. 2020).

Based on these results and other studies (Drewnick et al. 2020), we recommend a three-layer mask consisting of two outer layers of a very flexible, tightly woven fabric and an inner layer consisting of a material designed to filter out particles. The inner layer could be a high efficiency particulate air (HEPA) filter, a MERV 14 or better filter (Azimi, Zhao and Stephens 2014), a good surgical mask, or a vacuum bag. This approach produces a good fitting mask with high performance in both directions. If the filter material is 60% efficient at the most penetrating particle size and the outer layers are 20% efficient (Figure 1), the mask would have a minimum efficiency of 74%. At a particle size of 1 μm, where filter materials can easily have an efficiency of 75% and common fabrics 40%, the overall efficiency would be greater than 90%.

## Conclusion

In this study, we evaluated the material filtration efficiency, inward protection efficiency, and outward protection efficiency of 10 masks and a face shield on a manikin, using NaCl aerosols over the size range of 0.04 μm to >100 μm. The vacuum bag performed best on all three metrics; it was capable of filtering out 60–96% of particles, and achieved an outward protection efficiency of 50%–95%% and an inward protection efficiency of 75%–96%% for particles of aerodynamic diameter 0.5 μm and greater. The thin acrylic performed worst, with a material filtration efficiency of <25% for particles at 0.1 μm and larger, and inward and outward protection efficiencies of <50%. The material filtration efficiency was generally positively correlated with either inward or outward protection effectiveness, but stiffer materials were an exception to this relationship as they did not fit as closely to the manikin. Factors including stiffness of the material, the way of wearing the mask (e.g., earloops vs. tied around the head), and material hydrophobicity affected the fit of the mask and thus its performance. Future studies may focus on the influence of material properties on the fit of the mask, and how the transmission of real viruses, including SARS-CoV-2, is altered by wearing the masks.

## Data Availability

Data are available upon request.

## Acknowledgments

Jin Pan was supported by an Edna B. Sussman Foundation fellowship. Virginia Tech’s Fralin Life Sciences Institute and Institute for Critical Technology and Applied Science provided additional support for this work. Elizabeth Cantando acquired SEM images. TSI Inc. generously loaned the Flow Focusing Monodisperse Aerosol Generator 1520 to the Marr lab. This work used shared facilities at the Virginia Tech National Center for Earth and Environmental Nanotechnology Infrastructure (NanoEarth), a member of the National Nanotechnology Coordinated Infrastructure (NNCI), supported by NSF (ECCS 1542100 and ECCS 2025151).

