## Supplemental Information for "Inward and outward effectiveness of cloth masks, a surgical mask, and a face shield"

### Supplementary information

To verify the influence of a neutralizer on the material filtration efficiency, we connected an aerosol neutralizer (soft X-ray type neutralizer XRC-05, HCT CO., Ltd, Republic of Korea) after the Collison nebulizer. We tested an N95 and found no significant difference in efficiency ($p$ > 0.05) whether the neutralizer was on or off, as determined by the Student's t-test (Figure S1). Figure S2 shows the results with microfiber cloth. The filtration efficiency when the neutralizer was off was slightly lower, but there was no significant difference over the whole range whether the neutralizer was on or off ($p$ > 0.05). We also tested the N95 with Kr-85 radioactive neutralizer (3012, TSI Inc., MN, USA) and found no significant difference ($p$ > 0.05) (Figure S3). Error bars represent standard deviations of duplicates.

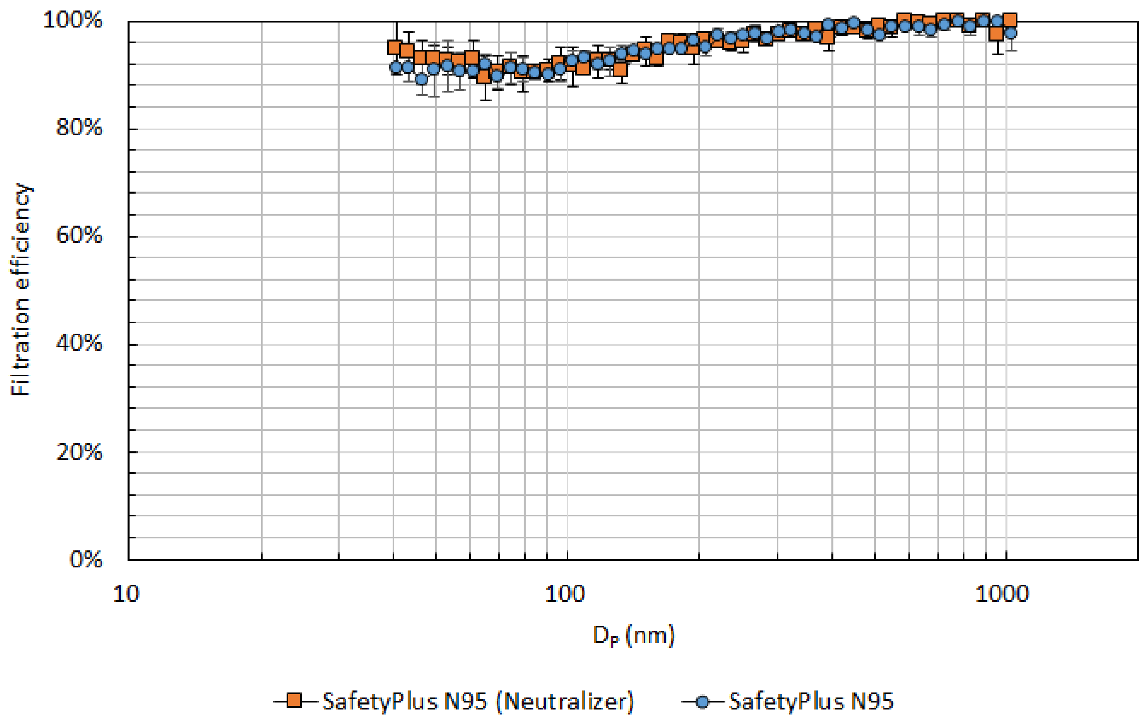


Figure S1. Material filtration efficiency of an N95 with and without a neutralizer (soft X-ray).

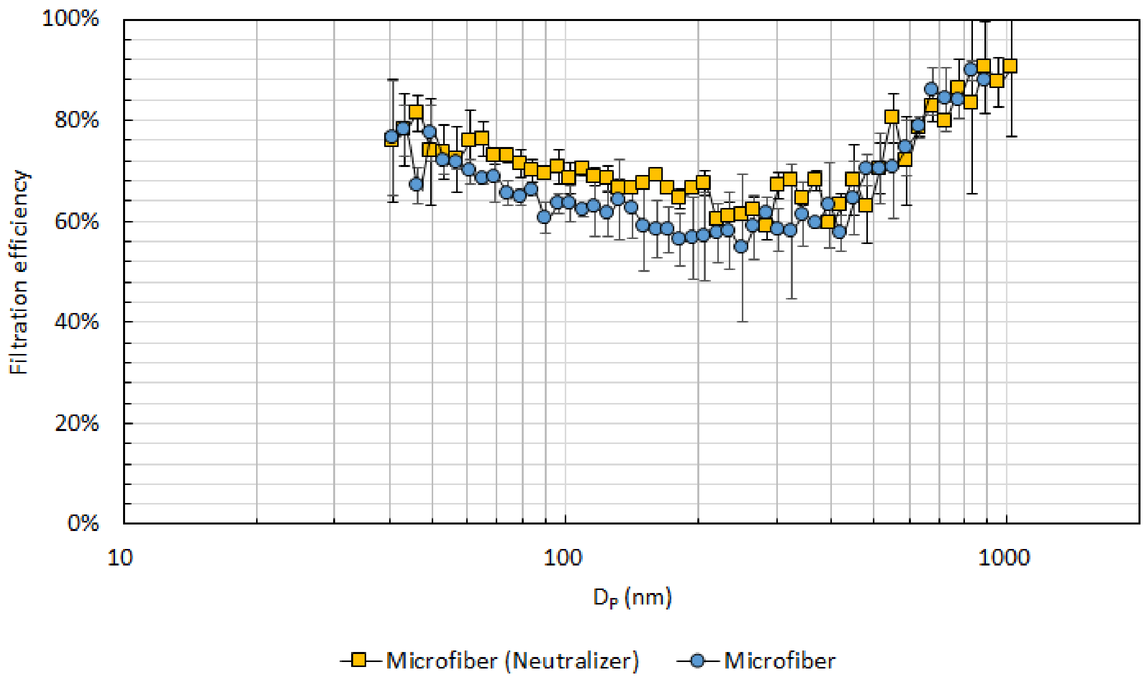


Figure S2. Material filtration efficiency of microfiber cloth with and without a neutralizer (soft X-ray).

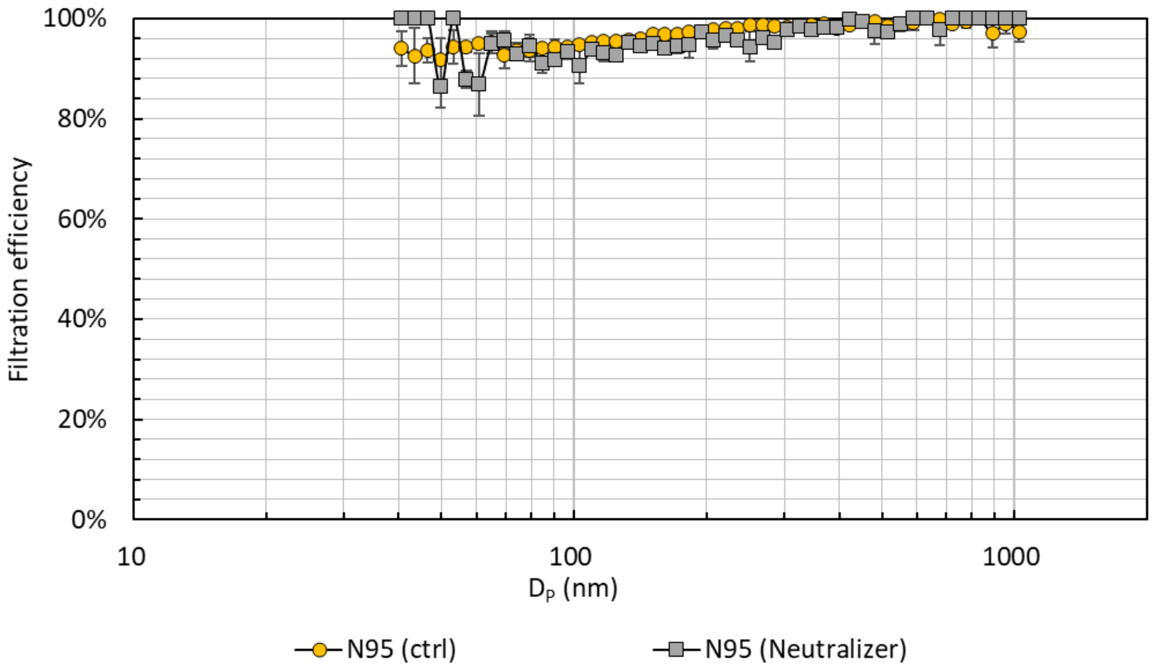


Figure S3. Material filtration efficiency of an N95 with and without a neutralizer (Kr-85).

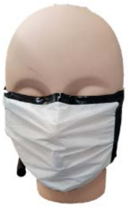

(a) Vacuum bag mask

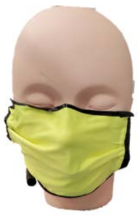

(b) Microfiber mask

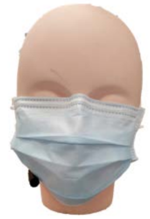

(c) Surgical mask

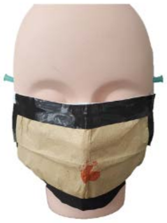

(d) Coffee filter mask

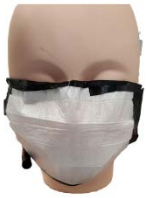

(e) MERV 12 filter mask

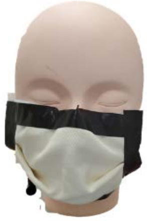

(f) Thin cotton mask

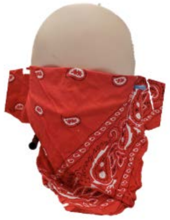

(g) Bandana

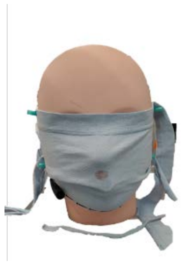

(h) CDC non-sewn

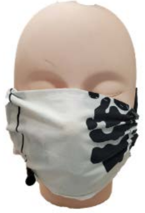

(i) CDC sewn

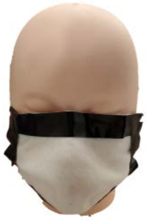

(j) Thin acrylic mask

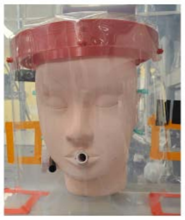

(k) Face shield

Figure S4. Masks on the manikin.

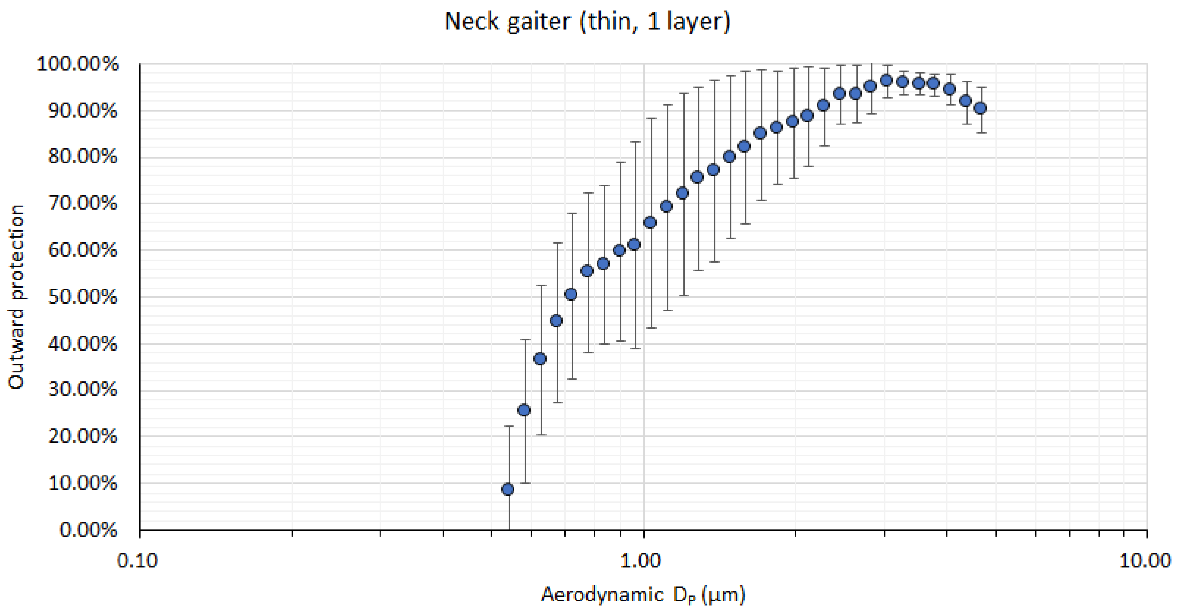


Figure S5. Outward protection efficiency of a thin, polyester neck gaiter (1 ply).

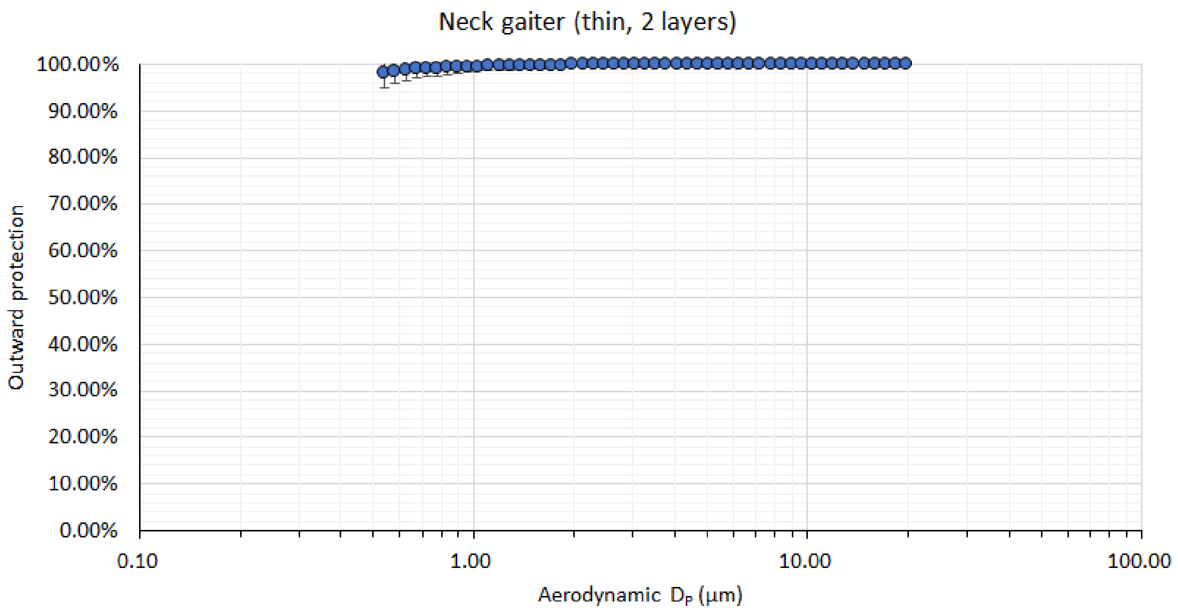


Figure S6. Outward protection efficiency of a thin, polyester neck gaiter (2 ply).

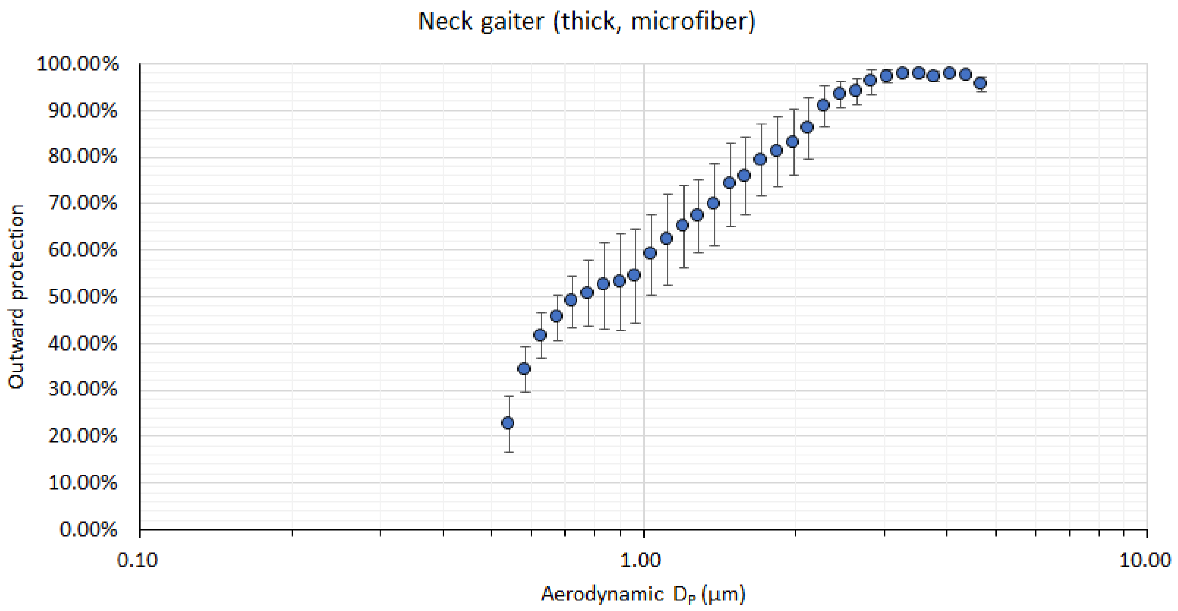


Figure S7. Outward protection efficiency of a double-layer, microfiber, polyester and elastane neck gaiter.
